# Evaluation of Viasure SARS-CoV-2 RT-qPCR kit (CerTest Biotec) using CDC FDA EUA RT-qPCR kit as a gold standard

**DOI:** 10.1101/2020.06.29.20131367

**Authors:** Byron Freire-Paspuel, Patricio Vega-Mariño, Alberto Velez, Paulina Castillo, Marilyn Cruz, Franklin Perez, Miguel Angel Garcia-Bereguiain

## Abstract

**Background:** Several RT-qPCR kits are available for SARS-CoV-2 diagnosis, some of them with Emergency Use Authorization (EUA) by FDA, but most of them lacking of proper evaluation studies due to covid19 emergency.

**Objective:** We evaluated Viasure RT-qPCR kit (CerTest Biotec, Spain) for SARS-CoV-2 diagnosis using CDC FDA EUA kit as gold standard.

**Results:** Although we found the lack of RNA quality control probe as the main limitation for Viasure kit, the sensitivity was up to 97.5% and specificity was 100%.

**Conclusions:** Viasure RT-qPCR kit is a reliable tool for SARS-CoV-2 diagnosis but improvement of an alternative RT-qPCR reaction for RNA extraction quality control as RNaseP is recommended.

## Background

Multiple in vitro RT-qPCR diagnosis kits are available on the market for the detection of SARS-CoV-2. Some of them have received emergency use authorization (EUA) from the U.S. Food & Drug Administration (FDA), while for others only limited report validations made by manufacturers are available. The CDC designed 2019-nCoV CDC EUA kit (IDT, USA) is based on N1 and N2 probes to detect SARS-CoV-2 that have received positive evaluation on recent reports (1–4), and and RNase P as an RNA extraction quality control. Among the commercial kits available in the market, Viasure SARS-CoV-2 RT-qPCR kit (CerTest Biotec; Spain) includes “ORF1ab” and “N” probes for SARS-CoV-2 detection. However, no probe for RNA extraction quality control is included but simply an “internal positive control” to guarantee that PCR reaction performs well. Viasure SARS-CoV-2 kit is made in Spain, one of the countries leading COVID19 cases and deaths worldwide, where it has been used for SARS-CoV-2 diagnosis. Also, it was recently authorized for SARS-CoV2 diagnosis in Ecuador. However, it is not included on the list of FDA EUA kits (5) and no evaluation study for Viasure SARS-CoV-2 RT-qPCR kit has been reported beyond the limited validation provided by manufacturer’s manual including only two SARS-CoV-2 positive samples (6).

### Objective

This study compared the performance in terms of sensitivity and specificity of Viasure SARS-CoV-2 and 2019-nCoV CDC EUA kits for SARS-CoV-2 RT-qPCR diagnosis from nasopharyngeal samples.

### Study design

104 clinical specimens (nasopharyngeal swabs collected on 0.5mL TE pH 8 buffer) were included on this study, coming from individuals selected for SARS-CoV-2 surveillance at “LabGal” (“Agencia de Regulación y Control de la Bioseguridad y Cuarentena para Galápagos”) in Galapagos Islands, starting on April 8th 2020. Also, four negative controls (TE pH 8 buffer) were included as control for carryover contamination.

## Results

104 samples were tested for SARS-CoV-2 following an adapted version of the CDC protocol (1,2) using CFX96 BioRad instrument and PureLink Viral RNA/DNA Mini Kit (Invitrogen, USA) as an alternate RNA extraction method; also, same RNA samples were tested using Viasure SARS-CoV-2 RT-qPCR kit following manufacturer’s manual (6). For the 2019-nCoV CDC EUA kit, 81 samples tested positive and 23 samples tested negative (Tables 1 and 2). Considering only true positive samples for Viasure SARS-CoV-2 kit (amplification for both N and Orf1ab probes), 74 samples tested positive and 30 samples were negative; but if we consider as positive not only true positives but also “presumptive positive” samples (only amplification of N probe), 79 samples tested positive and 25 were negative (Tables 1 and 2). The range of Ct values obtained for 2019-nCoV CDC EUA were 22.61-38.58 and 24.02-41.7 for N1 and N2, respectively (Table 2). The range of Ct values obtained for Viasure SARS-CoV-2 kit were 22.50-38.00 and 22.67-40.03 for N and Orf1ab, respectively (Table 2). The viral loads detailed on Table 2 were calculated running a calibration curve with 2019-nCoV N positive control (IDT, USA), with a detection limit below 1 viral RNA copy. Viral loads (copies/uL) values for 2019-nCoV CDC EUA kit positive/Viasure SARS-CoV-2 kit negative samples were 2 (sample ID 999c) and 3,8 (sample ID 1603); viral loads (copies/uL) values for 2019-nCoV CDC EUA kit positive/Viasure SARS-CoV-2 kit presumptive positive samples were 4.900 (sample ID 448), 8,8 (sample ID 772), 4,36 (sample ID 1575), 2,96 (sample ID 1581) and 0,71 (sample ID 782) (See Table 2).

**Table 1.** **Performance of Viasure SARS-CoV-2 compared to 2019-nCoV CDC EUA for RT-qPCR SARS-CoV-2 diagnosis (% values: sensitivity). Presumptive positive samples only amplified for N probe with Viasure SARS-CoV-2 kit**.

|  | **Viasure Positive** | **Viasure positive + presumptive positive** | **Viasure Negative** | **Total** |
| --- | --- | --- | --- | --- |
| **CDC Positive** | 74 (91.4%) | 79 (97.5%) | 2 | 81 |
| **CDC Negative** | 0 | 0 | 23 | 23 |
| **Total** | 74 | 79 | 25 | 104 |

**Table 2.**
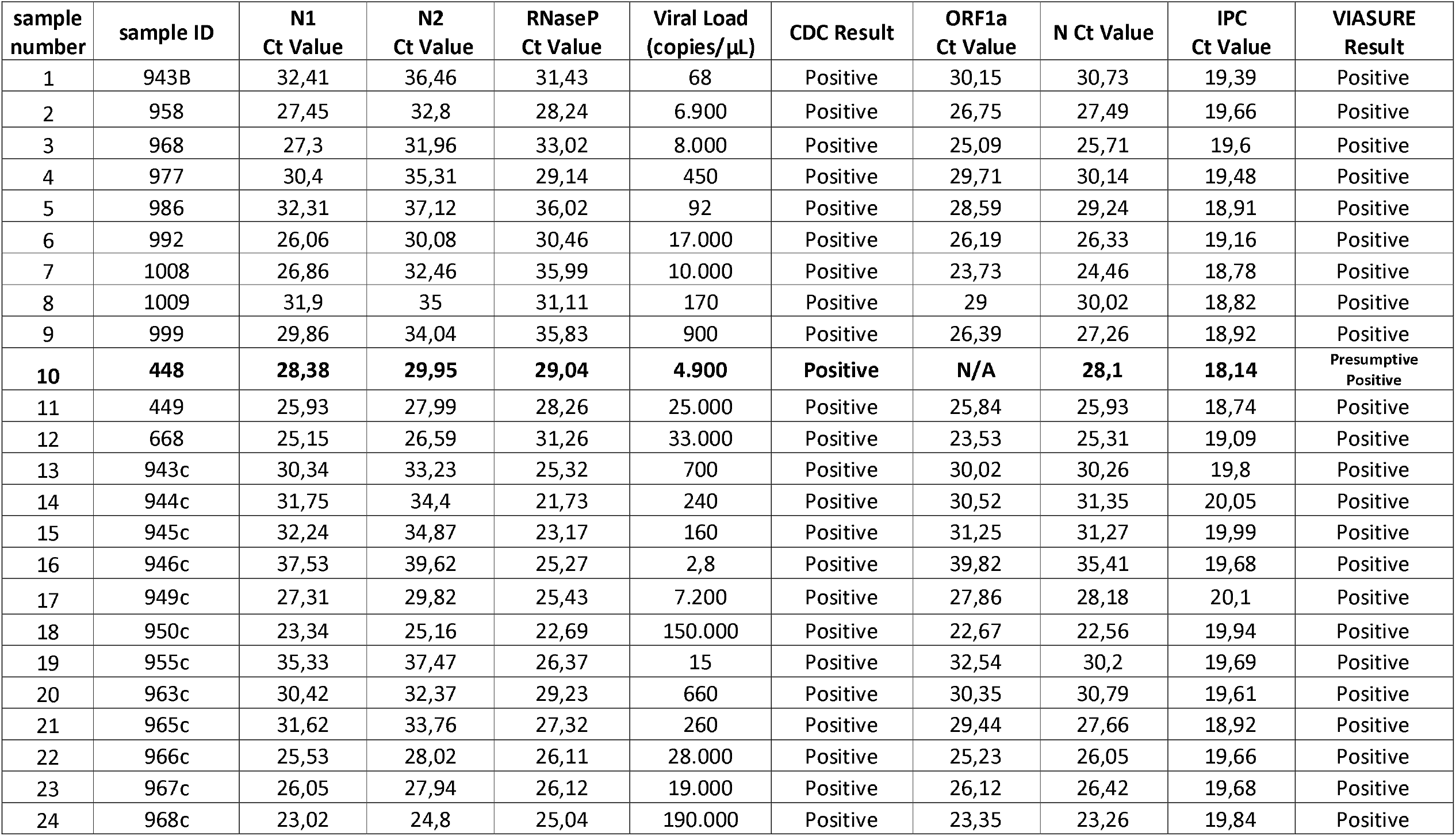
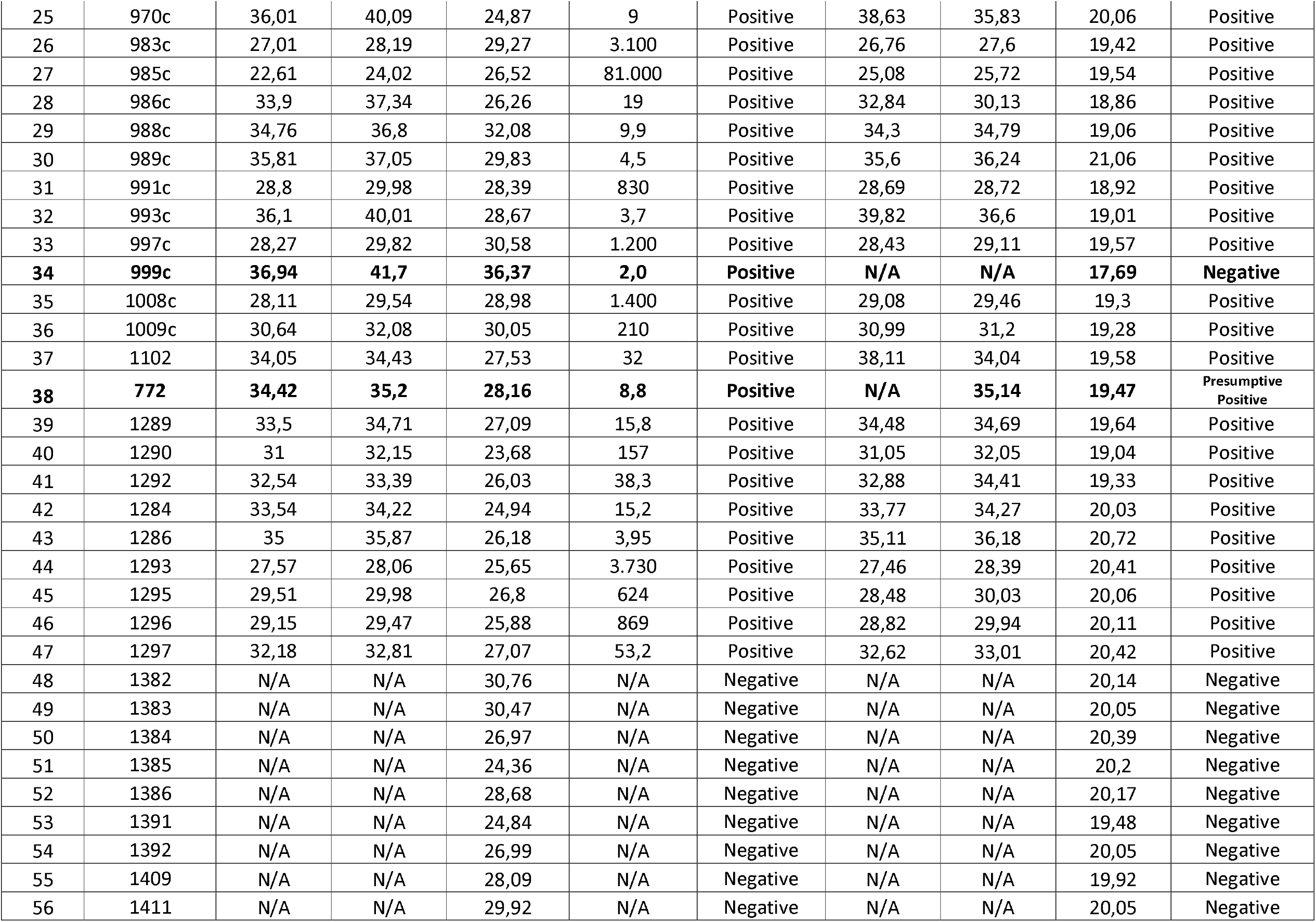
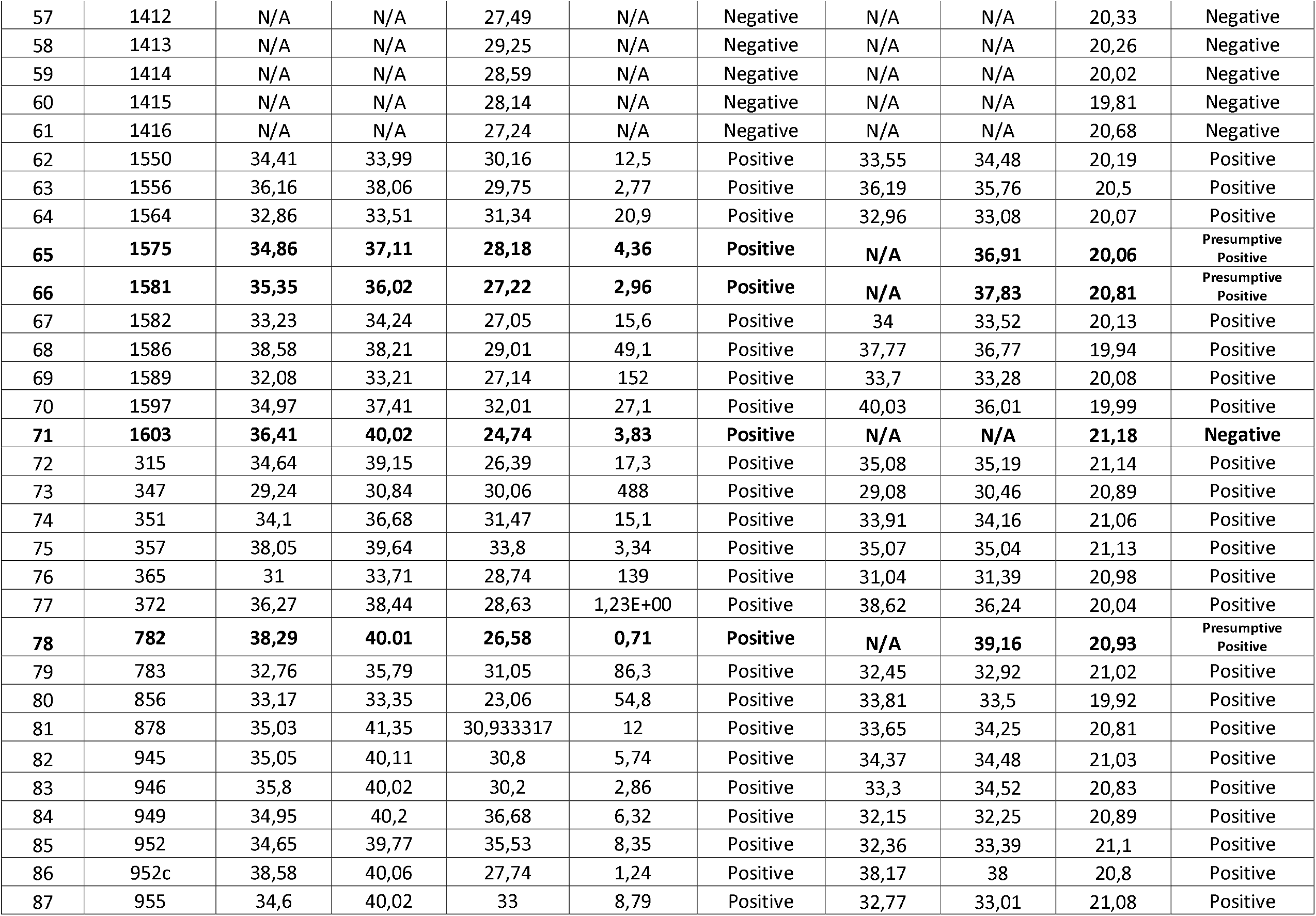
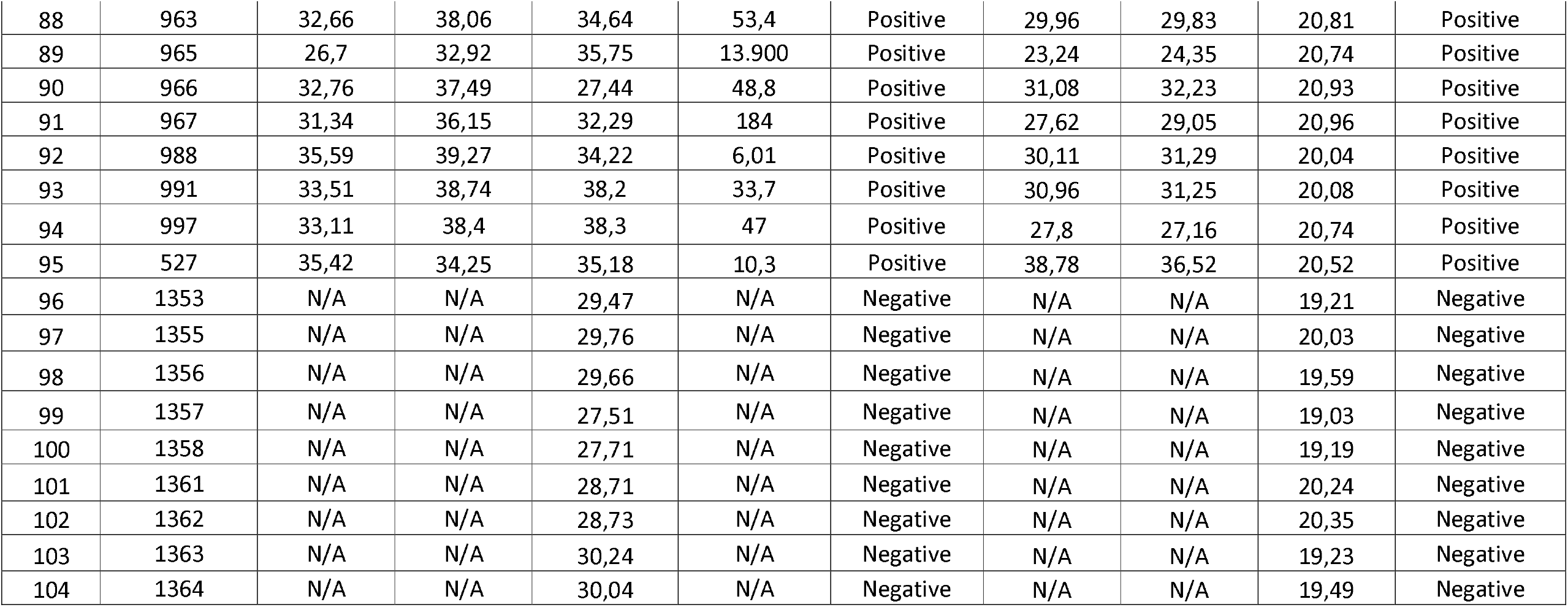
**Ct Values for nCoV-2019 CDC EUA kit and Viasure SARS-CoV-2 RT-qPCR kits for all the samples included on the study. RNaseP is the RNA extraction quality control for nCoV-2019 CDC EUA kit. IPC is the internal positive control for Viasure SARS-CoV-2 kit. Samples in bold were positive for nCoV-2019 CD EUA kit but either negative or presuntive positive for Viasure SARS-CoV-2 kit (N/A means not amplified)**.

| sample number | sample ID | N1 Ct Value | N2 Ct Value | RNaseP Ct Value | Viral Load (copies/μL) | CDC Result | ORF1a Ct Value | N Ct Value | IPC Ct Value | VIASURE Result |
| --- | --- | --- | --- | --- | --- | --- | --- | --- | --- | --- |
| 1 | 943B | 32,41 | 36,46 | 31,43 | 68 | Positive | 30,15 | 30,73 | 19,39 | Positive |
| 2 | 958 | 27,45 | 32,8 | 28,24 | 6.900 | Positive | 26,75 | 27,49 | 19,66 | Positive |
| 3 | 968 | 27,3 | 31,96 | 33,02 | 8.000 | Positive | 25,09 | 25,71 | 19,6 | Positive |
| 4 | 977 | 30,4 | 35,31 | 29,14 | 450 | Positive | 29,71 | 30,14 | 19,48 | Positive |
| 5 | 986 | 32,31 | 37,12 | 36,02 | 92 | Positive | 28,59 | 29,24 | 18,91 | Positive |
| 6 | 992 | 26,06 | 30,08 | 30,46 | 17.000 | Positive | 26,19 | 26,33 | 19,16 | Positive |
| 7 | 1008 | 26,86 | 32,46 | 35,99 | 10.000 | Positive | 23,73 | 24,46 | 18,78 | Positive |
| 8 | 1009 | 31,9 | 35 | 31,11 | 170 | Positive | 29 | 30,02 | 18,82 | Positive |
| 9 | 999 | 29,86 | 34,04 | 35,83 | 900 | Positive | 26,39 | 27,26 | 18,92 | Positive |
| **10** | **448** | **28,38** | **29,95** | **29,04** | **4.900** | **Positive** | **N/A** | **28,1** | **18,14** | Presumptive Positive |
| 11 | 449 | 25,93 | 27,99 | 28,26 | 25.000 | Positive | 25,84 | 25,93 | 18,74 | Positive |
| 12 | 668 | 25,15 | 26,59 | 31,26 | 33.000 | Positive | 23,53 | 25,31 | 19,09 | Positive |
| 13 | 943c | 30,34 | 33,23 | 25,32 | 700 | Positive | 30,02 | 30,26 | 19,8 | Positive |
| 14 | 944c | 31,75 | 34,4 | 21,73 | 240 | Positive | 30,52 | 31,35 | 20,05 | Positive |
| 15 | 945c | 32,24 | 34,87 | 23,17 | 160 | Positive | 31,25 | 31,27 | 19,99 | Positive |
| 16 | 946c | 37,53 | 39,62 | 25,27 | 2,8 | Positive | 39,82 | 35,41 | 19,68 | Positive |
| 17 | 949c | 27,31 | 29,82 | 25,43 | 7.200 | Positive | 27,86 | 28,18 | 20,1 | Positive |
| 18 | 950c | 23,34 | 25,16 | 22,69 | 150.000 | Positive | 22,67 | 22,56 | 19,94 | Positive |
| 19 | 955c | 35,33 | 37,47 | 26,37 | 15 | Positive | 32,54 | 30,2 | 19,69 | Positive |
| 20 | 963c | 30,42 | 32,37 | 29,23 | 660 | Positive | 30,35 | 30,79 | 19,61 | Positive |
| 21 | 965c | 31,62 | 33,76 | 27,32 | 260 | Positive | 29,44 | 27,66 | 18,92 | Positive |
| 22 | 966c | 25,53 | 28,02 | 26,11 | 28.000 | Positive | 25,23 | 26,05 | 19,66 | Positive |
| 23 | 967c | 26,05 | 27,94 | 26,12 | 19.000 | Positive | 26,12 | 26,42 | 19,68 | Positive |
| 24 | 968c | 23,02 | 24,8 | 25,04 | 190.000 | Positive | 23,35 | 23,26 | 19,84 | Positive |
| 25 | 970c | 36,01 | 40,09 | 24,87 | 9 | Positive | 38,63 | 35,83 | 20,06 | Positive |
| 26 | 983c | 27,01 | 28,19 | 29,27 | 3.100 | Positive | 26,76 | 27,6 | 19,42 | Positive |
| 27 | 985c | 22,61 | 24,02 | 26,52 | 81.000 | Positive | 25,08 | 25,72 | 19,54 | Positive |
| 28 | 986c | 33,9 | 37,34 | 26,26 | 19 | Positive | 32,84 | 30,13 | 18,86 | Positive |
| 29 | 988c | 34,76 | 36,8 | 32,08 | 9,9 | Positive | 34,3 | 34,79 | 19,06 | Positive |
| 30 | 989c | 35,81 | 37,05 | 29,83 | 4,5 | Positive | 35,6 | 36,24 | 21,06 | Positive |
| 31 | 991c | 28,8 | 29,98 | 28,39 | 830 | Positive | 28,69 | 28,72 | 18,92 | Positive |
| 32 | 993c | 36,1 | 40,01 | 28,67 | 3,7 | Positive | 39,82 | 36,6 | 19,01 | Positive |
| 33 | 997c | 28,27 | 29,82 | 30,58 | 1.200 | Positive | 28,43 | 29,11 | 19,57 | Positive |
| **34** | **999c** | **36,94** | **41,7** | **36,37** | **2,0** | **Positive** | **N/A** | **N/A** | **17,69** | **Negative** |
| 35 | 1008c | 28,11 | 29,54 | 28,98 | 1.400 | Positive | 29,08 | 29,46 | 19,3 | Positive |
| 36 | 1009c | 30,64 | 32,08 | 30,05 | 210 | Positive | 30,99 | 31,2 | 19,28 | Positive |
| 37 | 1102 | 34,05 | 34,43 | 27,53 | 32 | Positive | 38,11 | 34,04 | 19,58 | Positive |
| **38** | **772** | **34,42** | **35,2** | **28,16** | **8,8** | **Positive** | **N/A** | **35,14** | **19,47** | **Presumptive Positive** |
| 39 | 1289 | 33,5 | 34,71 | 27,09 | 15,8 | Positive | 34,48 | 34,69 | 19,64 | Positive |
| 40 | 1290 | 31 | 32,15 | 23,68 | 157 | Positive | 31,05 | 32,05 | 19,04 | Positive |
| 41 | 1292 | 32,54 | 33,39 | 26,03 | 38,3 | Positive | 32,88 | 34,41 | 19,33 | Positive |
| 42 | 1284 | 33,54 | 34,22 | 24,94 | 15,2 | Positive | 33,77 | 34,27 | 20,03 | Positive |
| 43 | 1286 | 35 | 35,87 | 26,18 | 3,95 | Positive | 35,11 | 36,18 | 20,72 | Positive |
| 44 | 1293 | 27,57 | 28,06 | 25,65 | 3.730 | Positive | 27,46 | 28,39 | 20,41 | Positive |
| 45 | 1295 | 29,51 | 29,98 | 26,8 | 624 | Positive | 28,48 | 30,03 | 20,06 | Positive |
| 46 | 1296 | 29,15 | 29,47 | 25,88 | 869 | Positive | 28,82 | 29,94 | 20,11 | Positive |
| 47 | 1297 | 32,18 | 32,81 | 27,07 | 53,2 | Positive | 32,62 | 33,01 | 20,42 | Positive |
| 48 | 1382 | N/A | N/A | 30,76 | N/A | Negative | N/A | N/A | 20,14 | Negative |
| 49 | 1383 | N/A | N/A | 30,47 | N/A | Negative | N/A | N/A | 20,05 | Negative |
| 50 | 1384 | N/A | N/A | 26,97 | N/A | Negative | N/A | N/A | 20,39 | Negative |
| 51 | 1385 | N/A | N/A | 24,36 | N/A | Negative | N/A | N/A | 20,2 | Negative |
| 52 | 1386 | N/A | N/A | 28,68 | N/A | Negative | N/A | N/A | 20,17 | Negative |
| 53 | 1391 | N/A | N/A | 24,84 | N/A | Negative | N/A | N/A | 19,48 | Negative |
| 54 | 1392 | N/A | N/A | 26,99 | N/A | Negative | N/A | N/A | 20,05 | Negative |
| 55 | 1409 | N/A | N/A | 28,09 | N/A | Negative | N/A | N/A | 19,92 | Negative |
| 56 | 1411 | N/A | N/A | 29,92 | N/A | Negative | N/A | N/A | 20,05 | Negative |
| 57 | 1412 | N/A | N/A | 27,49 | N/A | Negative | N/A | N/A | 20,33 | Negative |
| 58 | 1413 | N/A | N/A | 29,25 | N/A | Negative | N/A | N/A | 20,26 | Negative |
| 59 | 1414 | N/A | N/A | 28,59 | N/A | Negative | N/A | N/A | 20,02 | Negative |
| 60 | 1415 | N/A | N/A | 28,14 | N/A | Negative | N/A | N/A | 19,81 | Negative |
| 61 | 1416 | N/A | N/A | 27,24 | N/A | Negative | N/A | N/A | 20,68 | Negative |
| 62 | 1550 | 34,41 | 33,99 | 30,16 | 12,5 | Positive | 33,55 | 34,48 | 20,19 | Positive |
| 63 | 1556 | 36,16 | 38,06 | 29,75 | 2,77 | Positive | 36,19 | 35,76 | 20,5 | Positive |
| 64 | 1564 | 32,86 | 33,51 | 31,34 | 20,9 | Positive | 32,96 | 33,08 | 20,07 | Positive |
| **65** | **1575** | **34,86** | **37,11** | **28,18** | **4,36** | **Positive** | **N/A** | **36,91** | **20,06** | Presumptive Positive |
| **66** | **1581** | **35,35** | **36,02** | **27,22** | **2,96** | **Positive** | **N/A** | **37,83** | **20,81** | Presumptive Positive |
| 67 | 1582 | 33,23 | 34,24 | 27,05 | 15,6 | Positive | 34 | 33,52 | 20,13 | Positive |
| 68 | 1586 | 38,58 | 38,21 | 29,01 | 49,1 | Positive | 37,77 | 36,77 | 19,94 | Positive |
| 69 | 1589 | 32,08 | 33,21 | 27,14 | 152 | Positive | 33,7 | 33,28 | 20,08 | Positive |
| 70 | 1597 | 34,97 | 37,41 | 32,01 | 27,1 | Positive | 40,03 | 36,01 | 19,99 | Positive |
| **71** | **1603** | **36,41** | **40,02** | **24,74** | **3,83** | **Positive** | **N/A** | **N/A** | **21,18** | **Negative** |
| 72 | 315 | 34,64 | 39,15 | 26,39 | 17,3 | Positive | 35,08 | 35,19 | 21,14 | Positive |
| 73 | 347 | 29,24 | 30,84 | 30,06 | 488 | Positive | 29,08 | 30,46 | 20,89 | Positive |
| 74 | 351 | 34,1 | 36,68 | 31,47 | 15,1 | Positive | 33,91 | 34,16 | 21,06 | Positive |
| 75 | 357 | 38,05 | 39,64 | 33,8 | 3,34 | Positive | 35,07 | 35,04 | 21,13 | Positive |
| 76 | 365 | 31 | 33,71 | 28,74 | 139 | Positive | 31,04 | 31,39 | 20,98 | Positive |
| 77 | 372 | 36,27 | 38,44 | 28,63 | 1,23E+00 | Positive | 38,62 | 36,24 | 20,04 | Positive |
| **78** | **782** | **38,29** | **40,01** | **26,58** | **0,71** | **Positive** | **N/A** | **39,16** | **20,93** | Presumptive Positive |
| 79 | 783 | 32,76 | 35,79 | 31,05 | 86,3 | Positive | 32,45 | 32,92 | 21,02 | Positive |
| 80 | 856 | 33,17 | 33,35 | 23,06 | 54,8 | Positive | 33,81 | 33,5 | 19,92 | Positive |
| 81 | 878 | 35,03 | 41,35 | 30,933317 | 12 | Positive | 33,65 | 34,25 | 20,81 | Positive |
| 82 | 945 | 35,05 | 40,11 | 30,8 | 5,74 | Positive | 34,37 | 34,48 | 21,03 | Positive |
| 83 | 946 | 35,8 | 40,02 | 30,2 | 2,86 | Positive | 33,3 | 34,52 | 20,83 | Positive |
| 84 | 949 | 34,95 | 40,2 | 36,68 | 6,32 | Positive | 32,15 | 32,25 | 20,89 | Positive |
| 85 | 952 | 34,65 | 39,77 | 35,53 | 8,35 | Positive | 32,36 | 33,39 | 21,1 | Positive |
| 86 | 952c | 38,58 | 40,06 | 27,74 | 1,24 | Positive | 38,17 | 38 | 20,8 | Positive |
| 87 | 955 | 34,6 | 40,02 | 33 | 8,79 | Positive | 32,77 | 33,01 | 21,08 | Positive |
| 88 | 963 | 32,66 | 38,06 | 34,64 | 53,4 | Positive | 29,96 | 29,83 | 20,81 | Positive |
| 89 | 965 | 26,7 | 32,92 | 35,75 | 13.900 | Positive | 23,24 | 24,35 | 20,74 | Positive |
| 90 | 966 | 32,76 | 37,49 | 27,44 | 48,8 | Positive | 31,08 | 32,23 | 20,93 | Positive |
| 91 | 967 | 31,34 | 36,15 | 32,29 | 184 | Positive | 27,62 | 29,05 | 20,96 | Positive |
| 92 | 988 | 35,59 | 39,27 | 34,22 | 6,01 | Positive | 30,11 | 31,29 | 20,04 | Positive |
| 93 | 991 | 33,51 | 38,74 | 38,2 | 33,7 | Positive | 30,96 | 31,25 | 20,08 | Positive |
| 94 | 997 | 33,11 | 38,4 | 38,3 | 47 | Positive | 27,8 | 27,16 | 20,74 | Positive |
| 95 | 527 | 35,42 | 34,25 | 35,18 | 10,3 | Positive | 38,78 | 36,52 | 20,52 | Positive |
| 96 | 1353 | N/A | N/A | 29,47 | N/A | Negative | N/A | N/A | 19,21 | Negative |
| 97 | 1355 | N/A | N/A | 29,76 | N/A | Negative | N/A | N/A | 20,03 | Negative |
| 98 | 1356 | N/A | N/A | 29,66 | N/A | Negative | N/A | N/A | 19,59 | Negative |
| 99 | 1357 | N/A | N/A | 27,51 | N/A | Negative | N/A | N/A | 19,03 | Negative |
| 100 | 1358 | N/A | N/A | 27,71 | N/A | Negative | N/A | N/A | 19,19 | Negative |
| 101 | 1361 | N/A | N/A | 28,71 | N/A | Negative | N/A | N/A | 20,24 | Negative |
| 102 | 1362 | N/A | N/A | 28,73 | N/A | Negative | N/A | N/A | 20,35 | Negative |
| 103 | 1363 | N/A | N/A | 30,24 | N/A | Negative | N/A | N/A | 19,23 | Negative |
| 104 | 1364 | N/A | N/A | 30,04 | N/A | Negative | N/A | N/A | 19,49 | Negative |

In summary, sensitivity for Viasure SARS-CoV-2 kit compared to 2019-nCoV CDC EUA was 91.4% if we considered only true positives, but up to 97.5% if we considered also presumptive positives as positive samples for SARS-CoV-2.

## Discussion

Although the main limitation of our study is the sample size (104 specimens), our results support that Viasure SARS-CoV-2 RT-qPCR kit had a great performance in terms on sensitivity and specificity compared to 2019-nCoV CDC EUA, with values up to 97.5 and 100%, respectively. According to Viasure SARS-CoV-2 kit manufacturer’s manual, when a sample only yields amplification for N probe but Orf1ab is negative, the result should be SARS-CoV-2 negative and a possible infection by other coronavirus must be considered (6). However, on our hands, all the Viasure SARS-CoV-2 N positive/Orf1ab negative were confirmed as SARS-CoV-2 positive by CDC protocol (1,2), and thus we referred to this samples as presumptive positive. Moreover, Viasure SARS-CoV-2 kit manufacturer’s manual set a limit of detection of 10 viral copies/uL for its product. With the only exception of sample 448 (viral load of 4.300 copies/uL), all the Viasure SARS-CoV-2 kit presumptive positive and negative samples yield viral loads below 10 copies/uL, confirming the expected limit of detection of the kit.

The lack of any probe for RNA extraction quality control like RNaseP is a limitation to be considered when using Viasure SARS-CoV-2 kit. Although in our experience, samples failing to yield any RNA after extraction are below 1%, an extra RT-qPCR reaction for a RNA extraction quality control probe would be recommended for laboratories starting SARS-CoV-2 diagnosis. On the other hand, Viasure SARS-CoV-2 kit is provided on precast format of 8 tubes strips containing a mix of enzymes, primers, buffer and nucleotides in stabilized format, so only rehydratation buffer and RNA samples have to be added. This format is a great advantage in terms of time saving and reduction on pippeting mistakes. Also, the manufacturer provided us upon request the concentration of the positive control included on Viasure SARS-CoV-2 kit (10.000 copies/uL). That means viral loads can be easy calculated without need of purchase any extra positive control, despite the manufacturer’s manual does not include instruction for viral load calculation (6).

Considering the worldwide high demand of reagents for SARS-CoV RT-qPCR diagnosis, supplies shortage is a fact, actually affecting harder to developing countries like Ecuador. Under this scenario, validation studies are helpful to guarantee the quality of the supplies in the market for every country in the world, as not necessary all the SARS-CoV-2 RT-qPCR kits show the performance indicated by manufacturer (7).

## Data Availability

All data available within the manuscript

## Ethical considerations

All samples have been submitted for routine patient care and diagnostics at Galapagos Islands. This study was authorized by “Comité de Operaciones Especiales Regional de Galápagos” that is leading board for Covid19 surveillance in Galapagos Islands. All data used in the current study was anonymized prior to being obtained by the authors.

## Authorship contribution statement

All authors contributed to study conceptualization, experimental procedures and revision and approval of final version of the manuscript.

Byron Freire-Paspuel and Miguel Angel García Bereguiain analyzed the data and wrote the manuscript.

## Acknowledgements

We thank the medical personnel from “Ministerio de Salud Pública” at Galapagos Islands and the staff from the “Agencia de Regulación y Control de la Bioseguridad y Cuarentena para Galápagos” for their support. We also thank Dr. Ronald Cedeño from OPS/WHO for his work during Covid 19 surveillance in Galapagos Islands. We specially thank Gabriel Iturralde, Oscar Espinosa and Dr Tannya Lozada from “Dirección General de Investigación de la Universidad de Las Américas”, and also the authorities from Universidad de Las Américas, for logistic support to make SARS-CoV-2 diagnosis possible in Galapagos Islands.

